# Single-cell RNA-seq reveals a persistent interferon signature in immune cells from Systemic lupus erythematosus patients with high versus low polygenic risk scores despite antimalarial treatment

**DOI:** 10.64898/2026.03.12.26348018

**Authors:** Ahmed Sayadi, Maija-Leena Eloranta, Nina Oparina, Marcus Wallgren, Elisabeth Skoglund, Martina Frodlund, Christopher Sjöwall, Lars Rönnblom, Dag Leonard

## Abstract

**Objectives:** Patients with Systemic lupus erythematosus (SLE) who carry a high genetic burden often experience more severe disease. To understand the molecular consequences of polygenic risk, we analyzed single-cell gene expression profiles in SLE patients stratified by genetic risk.

**Methods:** Single-cell RNA sequencing (scRNA-seq) was performed on fresh peripheral blood mononuclear cells (PBMCs) from 16 female SLE patients, stratified by a weighted polygenic risk score (PRS), and 6 healthy controls (HCs). All patients were in low disease activity (LLDAS) and treated with antimalarials only. We assessed differential gene expression, interferon (IFN) signatures, transcription factor (TF) activity, and pathway enrichment across groups.

**Results:** Patients with High-PRS had significantly elevated IFN scores compared to HCs (p<0.001), whereas no significant difference was observed between Low-PRS patients and HCs (p>0.05) This pattern held across multiple immune cell types, including T cells, NK cells, and monocytes. Notable genes with increased expression in High-PRS patients included *ISG15* and *USP18* in plasmacytoid dendritic cells (pDCs), and *IFI27* and *RSAD2* in monocytes. IFN-related pathways were enriched in pDCs and monocytes in High-PRS patients, and only in monocytes in Low-PRS patients. TF analysis identified IRF7 and BATF3 as key candidate regulators in High-PRS of both cell types.

**Conclusions:** High polygenic risk in SLE is associated with persistent activation of IFN signaling pathways, indicating that antimalarial treatment alone is insufficient to fully suppress IFN activity, even during remission or low disease activity.

## 1. INTRODUCTION

Systemic lupus erythematosus (SLE) is a chronic, multisystem autoimmune disease characterized by the production of autoantibodies, immune complex deposition, and sustained activation of the type I interferon (IFN I) system[1, 2]. The clinical manifestations of SLE are highly heterogeneous, involving organs such as the kidneys, skin, joints, and central nervous system[3]. Despite the introduction of several targeted therapies in recent years, a substantial proportion of patients still develop irreversible organ damage and experience reduced survival[4–6].

Genetic susceptibility plays a fundamental role in SLE pathogenesis, supported by a concordance rate of approximately 50% in monozygotic twins[7]. Genome-wide association studies (GWAS) have now identified more than 300 loci associated with SLE at genome-wide significance (p < 5.0 × 10⁻⁸)[8, 9]. Most of these risk variants are common and confer modest individual effects, reflecting the polygenic nature of the disease[10]. Polygenic risk scores (PRSs) provide a means to quantify this cumulative genetic burden[11]. We have previously developed a weighted PRS for SLE and demonstrated that a higher genetic risk is associated with more severe clinical manifestations, increased organ damage, and reduced survival[12].

Single-cell RNA sequencing (scRNA-seq) enables unbiased characterization of gene expression at cellular resolution and has substantially advanced understanding of immune dysregulation in SLE. Studies of peripheral blood mononuclear cells (PBMCs) have revealed activation of the type I IFN system across multiple cell subsets including monocytes and dendritic cells[13]. Moreover, expanded populations of IFN-stimulated monocytes and T cells have been observed in patients with active disease[14].

In the present study, we stratify patients with SLE who are in Lupus Low Disease Activity State (LLDAS) and receiving antimalarial therapy alone according to their polygenic risk. Using scRNA-seq of PBMCs, we investigate how cumulative genetic susceptibility influences immune cell-specific transcriptional programs[15]. By linking polygenic risk to single-cell gene expression profiles, this study aims to provide mechanistic insight into how inherited genetic burden shapes immune dysregulation in SLE.

## 2. PATIENTS AND METHODS

### 2.1. Patients and controls

In total, 16 female patients fulfilling ≥4 of the American College of Rheumatology (ACR) classification criteria for SLE were recruited from Uppsala University Hospital and Linköping University Hospital, along with six age- and sex-matched healthy controls [16, 17]. SLE patients were selected based on their PRS for SLE, as defined by Reid *et al.* [12]. From a cohort of 353 patients with available PRS data, individuals at the highest and lowest ends of the PRS distribution were invited to participate. Additional inclusion criteria for patients were: female sex, Caucasian ethnicity, and achievement of LLDAS as assessed by a rheumatologist[15]. All participants were required to have a SLEDAI-2K score ≤4, stable treatment with antimalarial therapy, and no ongoing corticosteroid or disease-modifying antirheumatic drug (DMARD) use[18]. Ultimately, nine patients with a high PRS (High-PRS) and seven with a low PRS (Low-PRS) were enrolled (Supplementary table S1). All patients were on Hydroxychloroquine (HCQ) except one patient in the Low-PRS group who was treated with chloroquine phosphate. Clinical data was retrieved from medical charts. The study was approved by the local ethics review authority (2009/013, 2016/155), and all participants gave their informed consent.

### 2.2. Genotyping and development of PRS

Patients were genotyped using the Illumina 200K Immunochip SNP array at Science for Life Laboratory in Uppsala, Sweden. For quality control procedures, see the supplementary methods file. A weighted PRS for SLE susceptibility was calculated for 353 patients, based on 57 established non-HLA SLE-associated single nucleotide variants (SNVs) identified at genome-wide significance in European populations, as previously described[12] (Supplementary table S2). For each SNV, the natural logarithm of the odds ratio for SLE susceptibility was multiplied by the number of risk alleles carried by each individual. The sum of these products constituted the PRS for each patient.

### 2.3. Single-Cell RNA Sequencing

Single-cell RNA libraries were generated by loading ∼10,000 freshly isolated PBMC per donor onto the Chromium Next GEM Single Cell 3’ v3.1 (Dual Index, 10x Genomics) device. The amplified cDNA libraries were sequenced using 2 × 150 bp reads on an Illumina NovaSeq 6000 S4 flow cell with v1.5 chemistry.

A total of 1.68 billion reads were generated across all samples. On average, each cell had 79,476 reads, with a median of 2,724 genes and 8,158 unique molecular identifiers (UMIs) detected per cell. Cell Ranger (v7.1) estimated approximately 9,636 high-quality cells per donor[19].

### 2.4. Preprocessing and Quality Control

Raw sequencing reads were processed using the Cell Ranger pipeline (v7.1, 10X Genomics) with default parameters, including alignment to the human reference genome (GRCh38) and generation of gene expression count matrices[19]. Multiple samples were aggregated into a single dataset with normalizing sequencing depth using the CellRanger aggr. function.

Dimensionality reduction and unsupervised clustering were initially performed using the Cell Ranger’s pipeline. Specifically, principal component analysis (PCA) was computed using the top variable genes to generate a low-dimensional representation of the data. A k-nearest neighbor graph was constructed based on PCAs results, and unsupervised clustering was carried out using graph-based Louvain community detection. Nonlinear dimensionality reduction for visualization was performed using t-distributed stochastic neighbor embedding (t-SNE) and Uniform Manifold Approximation and Projection (UMAP).

Further downstream analysis was conducted in R (v4.3.0) using the Seurat package (v5.3.0)[20]. Cells were filtered based on quality control metrics: cells were retained if they had at least 200 detected genes, more than three total UMI counts, and less than 20% mitochondrial gene expression. Normalization, variable feature selection, and scaling were performed following Seurat’s standard workflow. Potential doublets were identified and removed using the scDblFinder package[21].

### 2.5. Cell annotation, differential Gene Expression, IFN score, pathway- and enrichment analysis

See the Supplementary methods file.

### 2.6. Measurement of IFN-α and HCQ in serum

IFN-α2 levels in serum were determined by using a single-molecule array SIMOA immunoassays IFN-α kit v.6 (Quanterix) with Lower Limit of Quantitation (LLOQ) of 0.03 pg/ml. HCQ concentration in sera was determined by liquid chromatography coupled to high resolution mass spectrometry[22]. LLOQ of the assay was 8.3 ng/ml.

### 2.7. Statistical analysis

Comparisons between groups were evaluated using Mann-Whitney U test. Fisher’s exact test was used for comparison of categorial variables. P values < 0.05 were considered significant.

## 3. RESULTS

### 3.1. Study population

To investigate the genetic contribution to immune cell expression profiles at single-cell level we selected patients from our SLE cohort with the highest (mean = 9.91 ± 0.63) and lowest (mean = 7.30 ± 0.59) PRS scores. No significant differences were observed between patient groups in disease duration, organ damage, or fulfillment of ACR classification criteria (Supplementary table S3). At the time of sampling, patients exhibited low disease activity, and patient-reported disease activity, assessed using the SLAQ questionnaire, did not differ significantly between the groups. The average weekly dose HCQ prescribed in the two groups were similar (High-PRS: 1.7 mg/week vs. Low-PRS: 1.8 mg/week, p=0.71). Similarly, neither serum HCQ (p=0.22) nor serum IFN-α (p=0.34) concentration differed significantly between the groups. Thus, we compared two groups receiving the similar antimalarial treatment in remission, but with divergent genetic risk profiles.

### 3.2. Genetic contribution to immune cell expression profiles at single-cell level

Following QC, on average 9,636 high-quality cells per donor remained for further analysis and there were no sample-type bias (Figure 1A). The final integrated dataset comprised 184,966 cells, with a mean of 2,724 genes detected per cell. Cell clusters within PBMCs were annotated as ten major immune cell populations, (Figure 1B) and the proportion of and recovery rates of the cell types were comparable across groups (Figures 1C-D). The distribution of immune cell populations was consistent with flow cytometric analysis of fresh PBMC (Supplementary table S4).

**Figure 1.**
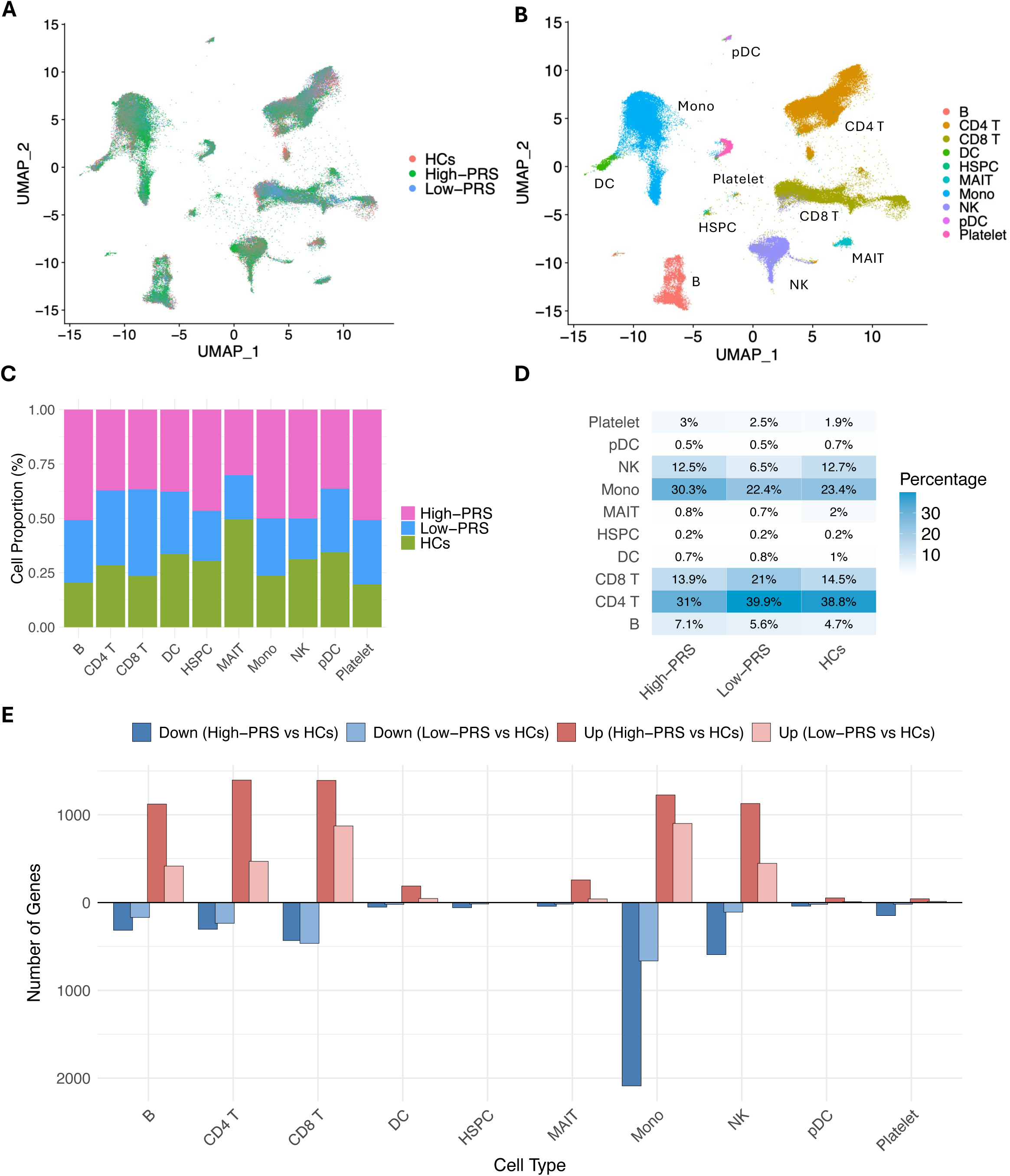
Single-cell RNA-seq analysis of peripheral blood mononuclear cells from SLE patients with high or low polygenic risk score (PRS) and healthy controls. (A) UMAP plot of cells colored by group: High-PRS, Low-PRS and Healthy controls (HCs), (B) UMAP plot of cells colored by cell type, including B cells, CD4 T cells, CD8 T cells, dendritic cells, hematopoietic stem and progenitor cells (HSPCs), mucosal-associated invariant T (MAIT) cells, monocytes, plasmacytoid dendritic cells (pDCs), and platelets. (C) Stacked bar plot showing the proportion of each cell type within each group, expressed as a percentage of normalized total cells per group. (D) Bar plot showing the percentage of each cell type within each group. (E) Bar plot showing the number of differentially expressed genes (upregulated in red, downregulated in blue) across different cell types when comparing SLE patients with High-PRS vs HCs and Low-PRS vs HCs.

We then conducted pairwise comparisons of differentially expressed genes (DEGs) across various immune cell types between SLE patients with high PRS or low PRS vs HCs. A greater number of DEGs were identified in more abundant cell populations, with the highest number observed in monocytes in the High-PRS vs. HCs comparison (Figure 1E).

### 3.3. Transcriptional differences between patients with SLE and healthy controls

To investigate the transcriptional differences between patients with SLE and HCs, we generated a volcano plot (Figure 2A). Several IFN-stimulated genes (ISGs), including sialic acid binding Ig-like lectin 1 (*SIGLEC1)*, IFN-induced protein 44-like (*IFI44L)* and IFN alpha-inducible protein 27 *(IFI27*), were significantly upregulated in patients, indicating a strong activation of the IFN signaling pathways. Pathway enrichment analysis using the Reactome database corroborated this finding, identifying, both type I IFN (α/β) and type II IFN (γ) signaling among the most significantly enriched pathways (Fig. 2B). To further quantify IFN activity, we calculated an IFN score, which was significantly elevated in patients compared to HCs (p = 0.0024), consistent with enhanced IFN pathway activation in SLE (Figure 2C).

**Figure 2.**
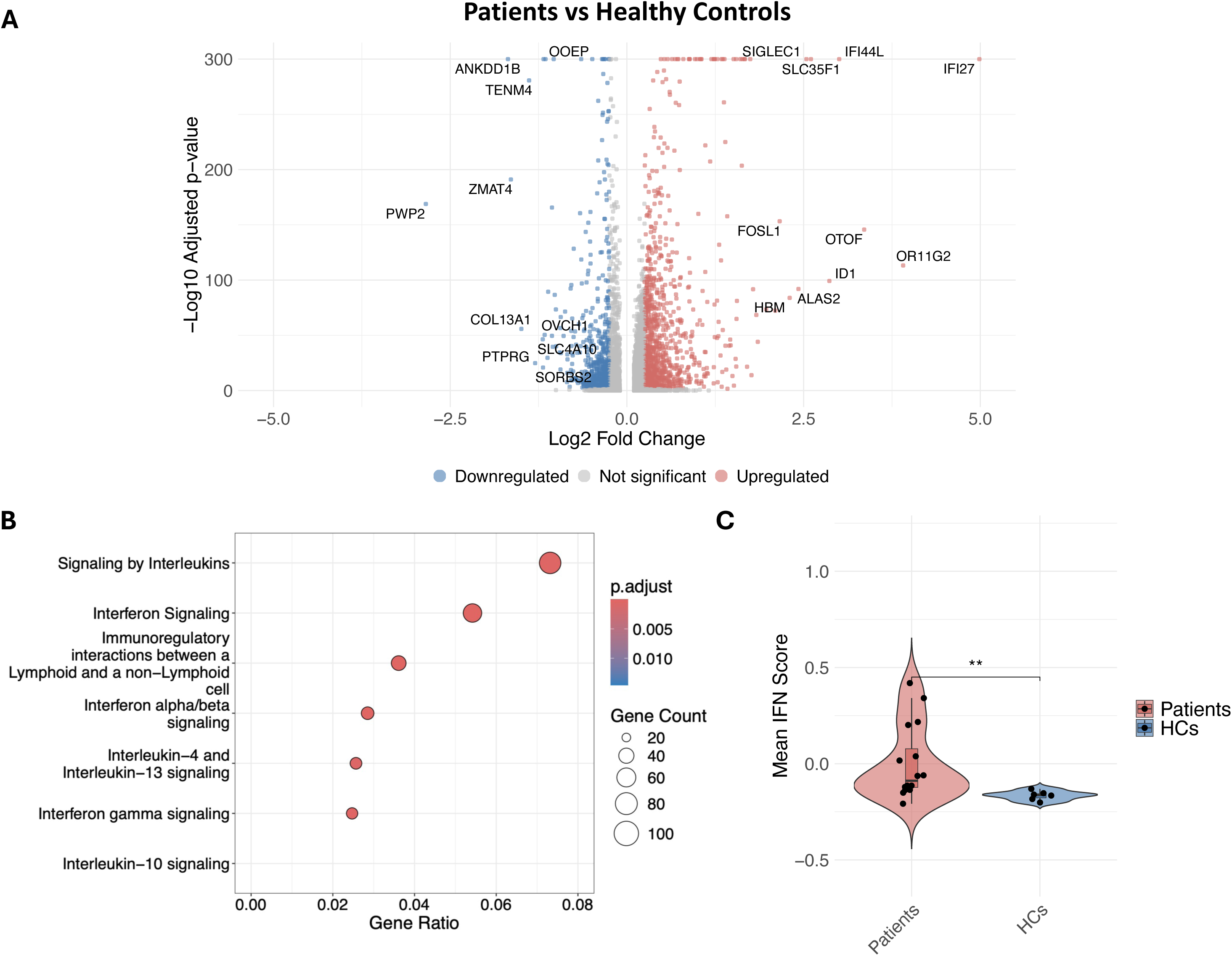
Transcriptional alterations and pathway enrichment in patients compared to controls. (A) Volcano plot of differentially expressed genes (DEGs) between patients with SLE and healthy controls (HCs). Genes with an adjusted p-value < 0.05 and absolute log₂ fold change > 0.25 are highlighted. (B) Dot plot showing the top enriched Reactome pathways among DEGs. Dot color represents statistical significance of the enrichment (−log₁₀ adjusted p-value), and dot size corresponds to the number of DEGs within each pathway. The x-axis indicates gene ratio, calculated as the number of DEGs in the pathway divided by the total number of genes annotated to that pathway. (C) Violin plots showing the distribution of interferon (IFN) response scores in patients and controls. Scores were computed per cell and averaged per sample. Statistical comparisons were performed using the Mann-Whitney U test.

### 3.4. Transcriptional activity of SLE cells stratified by genetic risk

To explore transcriptional variation across groups, we analyzed DEGs in four key comparisons: all patients vs. HCs, High-PRS vs. HCs, Low-PRS vs. HCs, and High-PRS vs. Low-PRS. To visualize the overlap between these gene sets, we generated an UpSet plot (Figure 3A) and Venn diagrams for all DEGs and ISG-specific DEGs (Figure 3B).

**Figure 3.**
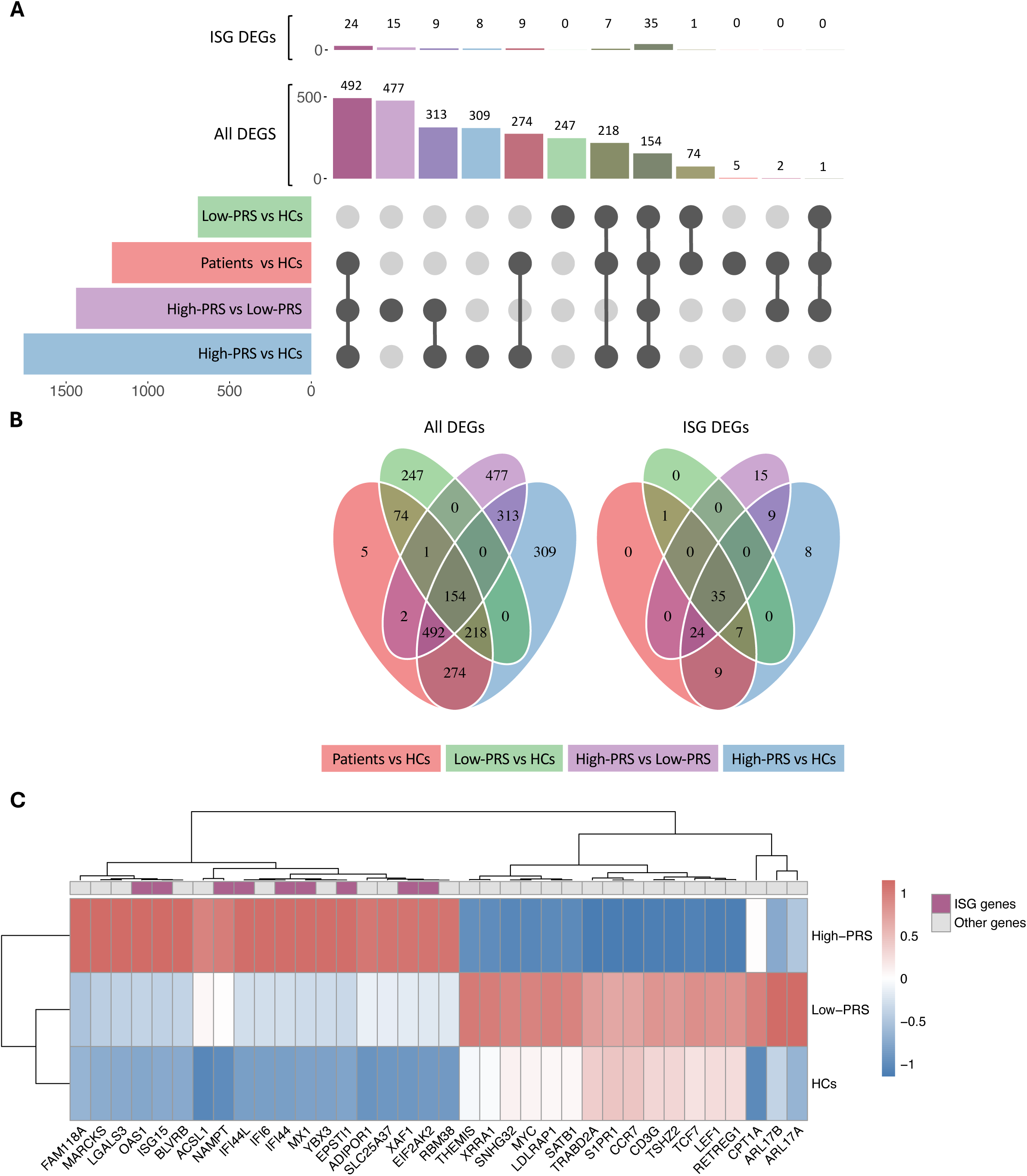
Differentially upregulated genes with overlap between SLE patients with High-PRS, Low-PRS and healthy controls. (A) UpSet plot and (B) Venn diagram illustrating the overlap of upregulated DEGs and IFN-stimulated genes (ISGs), defined by the Hallmark type I/II set, among all patients vs. HCs, High-PRS vs. HC, Low-PRS vs. HCs and High-PRS vs. Low-PRS. (C) Heatmap of the top differentially expressed genes across three groups: High-PRS, Low-PRS, and HCs. ISGs are shown in purple. Expression values are scaled by gene, and hierarchical clustering was applied to both genes and samples.

Across all comparisons, 154 genes were consistently upregulated in patients, indicating a shared core transcriptional program regardless of PRS. The High-PRS group exhibited the largest number of upregulated genes (1,760 genes) compared with HCs (Figure 3B). In contrast, the Low-PRS vs. HCs comparison revealed 694 upregulated genes, reflecting a more limited transcriptional response. Notably, a distinct set of 313 DEGs, including Complement C3b/C4b Receptor Like (*CR1L*, ≥2.4-fold change) and Basic Leucine Zipper ATF-Like Transcription Factor 2 (*BATF2*, ≥2.0-fold change), was exclusively shared between the High-PRS vs. HCs and the High-PRS vs Low-PRS comparisons. Together, these findings underscore the relative transcriptional quiescence of the Low-PRS group and highlight the influence of genetic background on the magnitude and nature of immune gene activation in SLE.

Focusing on ISGs, we identified 35 ISGs among the 154 DEGs shared across all comparisons, representing approximately 23% of the commonly upregulated genes (Figure 3B). An additional 33 ISGs were differentially expressed in both the High-PRS vs. HCs and High-PRS vs. Low-PRS comparisons.

To further characterize the expression dynamics across groups, we generated a heatmap of the top DEGs, including ISGs (Figure 3C). These analyses revealed that the genes highly upregulated in the High-PRS group, including 9 IFN-related DEGs, were less upregulated or even downregulated in the Low-PRS and HC groups. Conversely, genes upregulated in the Low-PRS or HC groups were often downregulated in the High-PRS group. This transcriptional divergence highlights a differential IFN response across PRS-stratified subgroups, with the High-PRS group showing a more pronounced activation of IFN-related genes/pathways.

### 3.5. IFN Score in Immune Cell Subsets Stratified by Polygenic Risk

Given the enrichment of both Type I and II IFN signaling pathways among the DEGs, we developed a composite IFN score using the top IFN-responsive genes annotated in the Hallmark gene set (Supplementary table S5). This score was significantly elevated in patients with high PRS compared to HCs (p=0.001; Figure 4A), while no significant difference was observed between Low-PRS individuals and HCs (Figure 4B). Cell-type-resolved analyses further revealed increased IFN score across multiple immune subsets in the High-PRS group (Figure 4C), with no significant changes detected in any subsets between Low-PRS individuals and HCs (Figure 4D). To dissect the specific contributions of type I and II IFNs, we computed separate scores for each subtype (Supplementary Figures S1-2). Both type I (p< 0.05) and type II IFN (p<0.01) scores were significantly higher in High-PRS vs HC comparisons, while no significant differences were observed between Low-PRS individuals and HCs.

**Figure 4.**
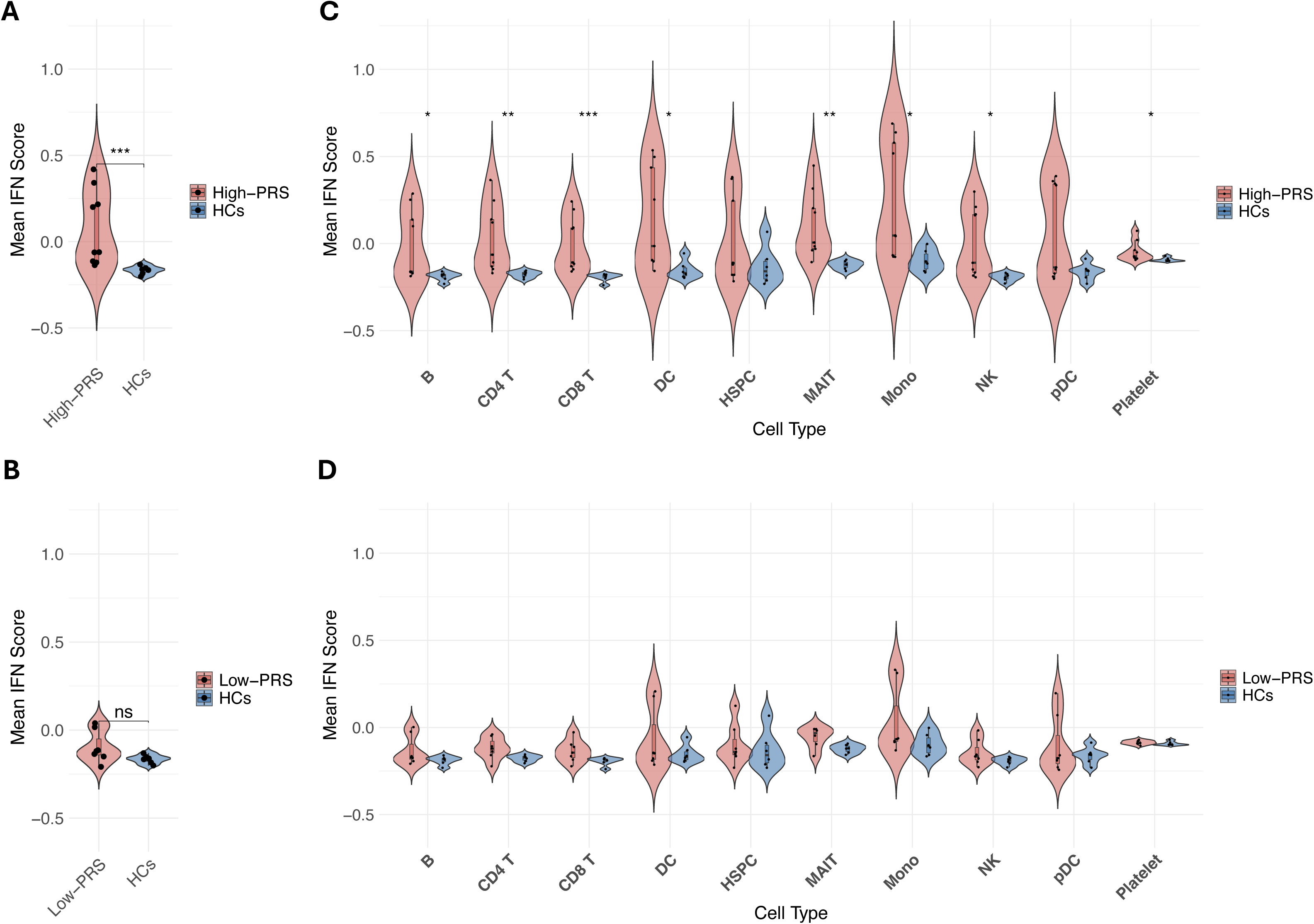
Cell type specific IFN score in SLE patients stratified by polygenic risk score. Violin plots depicting the distribution of interferon (IFN) scores per sample across all annotated cell types. (A) Comparison of High-PRS versus HCs and (B) Low-PRS versus HCs. The scores were calculated per cell and averaged per sample. (C, D) Violin plots showing IFN response scores stratified by cell types for (C) High-PRS vs HCs and (D) Low-PRS vs HCs. Statistical significance was evaluated using the Mann-Whitney U test.

Similarly, cell-type–resolved analyses showed significantly elevated type I and II IFN scores across most immune cell subsets in the High-PRS group, but not in the Low-PRS group. An exception was noted, for the type I IFN score in B cells, which was also elevated in Low-PRS individuals. Together, these findings suggest that high polygenic risk for SLE is associated with increased basal IFN pathway activation across diverse immune cell types.

### 3.6. Interferon-Stimulated Gene Expression Across Cell Subsets

To further investigate the DEGs contributing to the IFN score, we analyzed ISG expression across immune cell subsets. Patients with high PRS exhibited robust upregulation of multiple ISGs in most cell types, including *IFI44L*, *MX1* and IFN-Stimulated Protein 15 (*ISG15*) (Supplementary Figures S3A-D; Supplementary table S6). In contrast, Low-PRS patients demonstrated weaker ISGs response, with moderate upregulation of selected genes such as *IFI44* in monocytes.

The largest fold changes between High- and Low-PRS groups, were in pDCs observed for *ISG15* and *USP18*, and in monocytes for *IFI27* and *RSAD2*. In HCs, expression of Interferon regulatory factor 7 (IRF7) was largely restricted to pDCs and conventional DCs, whereas in the High-PRS group, *IRF7* expression extended to NK cells, B cells, and monocytes (Supplementary figures S3A-D). Significant differential expression of *IRF7* between High-and Low-PRS groups was observed in monocytes, but not in pDCs. Taken together, these results indicate that elevated polygenic risk is associated with broad dysregulation of IFN-responsive genes across multiple immune cell types.

#### 3.7.1. Pathway Enrichment Analysis in pDCs and Monocytes Stratified by Polygenic Risk

We next performed pathway enriched analysis on pDCs and monocytes, important producers of type I IFNs, to explore functional differences associated with polygenic risk (Figure 5).

**Figure 5.**
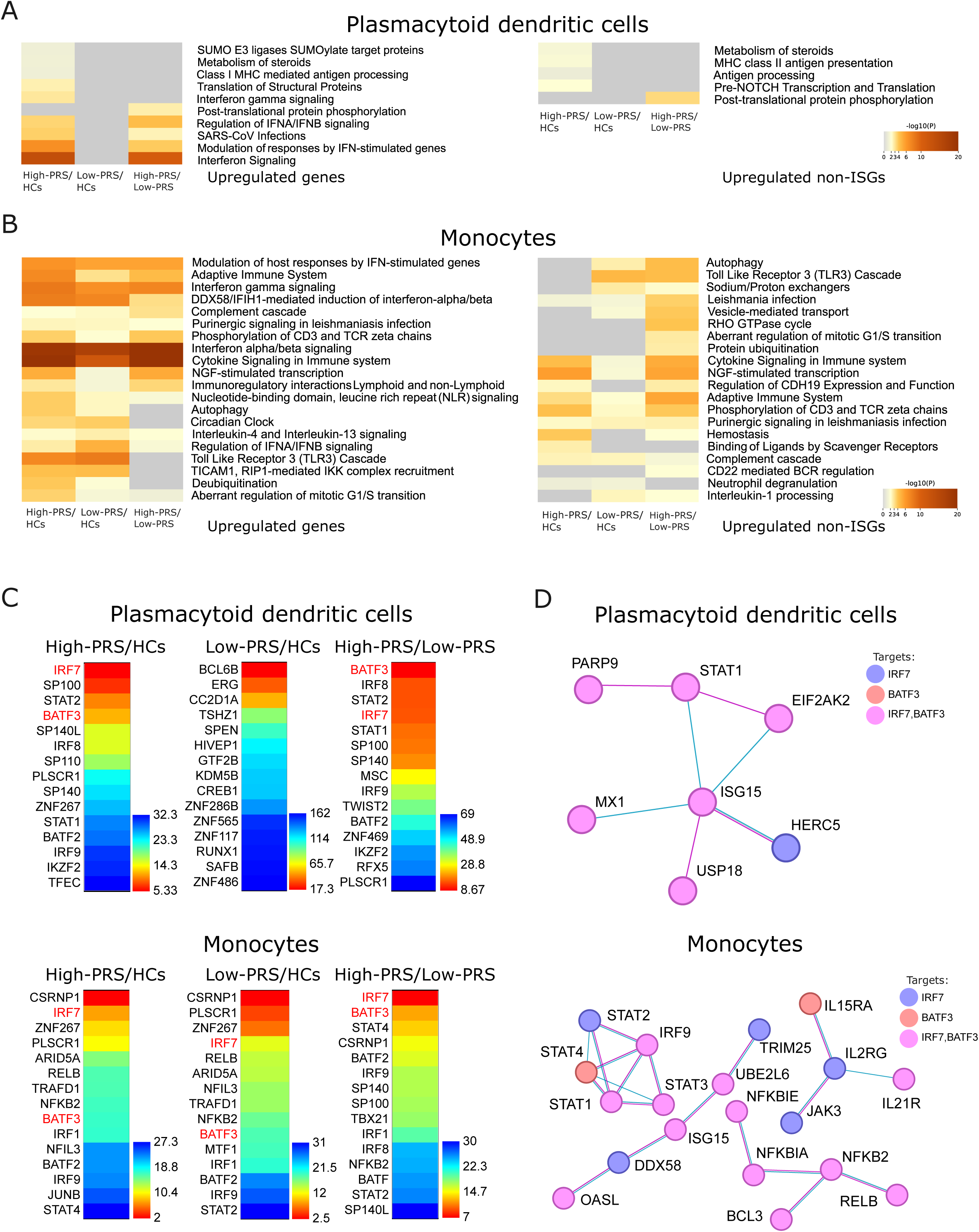
Functional enrichment analysis of genes upregulated in pDCs and monocytes from PRS-stratified patients with SLE compared with healthy controls. Heatmaps displaying pathway enrichment (log-transformed adjusted-values) for genes upregulated in (A) plasmacytoid dendritic cells (pDCs) and (B) monocytes. Comparisons include: High-PRS SLE patients vs. HCs, High-PRS vs Low-PRS patients, and Low-PRS vs. HCs. “Upregulated genes” refers to all upregulated DEGs; “Upregulated non-ISGs” excludes DEGs annotated as Hallmark ISGs. (C) Heatmaps showing transcription factor (TF) enrichment for the same comparisons using ChEA[47], identifying potential upstream regulators of the upregulated DEGs. The top 15 enriched TFs are displayed for each comparison (D) STRING-derived core physical protein-protein interaction networks for DEGs upregulated in pDCs and monocytes from High-PRS patients compared with HCs. The number of connecting lines indicates the number of supporting STRING evidence sources. Circle colors represent predicted target genes regulated by key TFs IRF7 and BATF3, as highlighted in (C).

In pDCs, immune-related pathways, including IFN signaling and regulation of IFN-α/β signaling, were significantly enriched in both High-PRS vs. HCs and High-PRS vs. Low-PRS comparisons. In contrast, no significant pathway enrichment was observed between Low-PRS and HCs (Figure 5A), indicating that type I IFN-related transcriptional activity in pDCs of HCQ treated patients, is primarily driven by individuals with higher genetic risk. To explore IFN-independent functional activity, we repeated the analysis after excluding ISGs, revealing a more limited set of enriched pathways in the High-PRS vs. HC comparison, most notably involving steroid metabolism and antigen processing. In the High-PRS vs. Low-PRS comparison, pathways such as post-translational protein phosphorylation emerged as enriched, suggesting alternative immune regulatory processes may also be influenced by genetic risk (Figure 5A).

In monocytes, enrichment of immune-related pathways, including IFN-α/β and IFN-γ signaling, was observed in High-PRS vs. HCs and High-PRS vs. Low-PRS, with a weaker enrichment observed also in the Low-PRS vs. HCs (Figure 5B). Notably, pathways such as NGF-stimulated transcription and phosphorylation of CD3 and TCR zeta chains were more highly enriched in High-PRS patients compared with both HCs and Low-PRS individuals. The Toll-like receptor 3 (TLR3) signaling pathway was specifically enriched in SLE monocytes relative to HCs but was not significantly different between High-PRS and Low-PRS groups. Following ISG exclusion, several pathways remained significantly enriched in monocytes, including adaptive immune system signaling, NGF signaling, and broader cytokine signaling pathways (Figure 5B), underscoring a persistent immune activation signature independent of ISG expression.

#### 3.7.2. Shared Transcriptional Regulators in pDCs and Monocytes

Shared functional patterns among upregulated genes in pDCs and monocytes pointed to common regulatory mechanisms, prompting transcription factor (TF) enrichment analysis using ChEA. Candidate TFs were prioritized based on binding specificity and cell-type expression. In pDCs, IRF7 ranked highest in the High-PRS vs. HCs comparison, while basic leucine zipper ATF-like transcription factor 3 (BATF3) emerged as the most enriched TF in the High-PRS vs. Low-PRS comparison (Figure 5C). In monocytes, both IRF7 and BATF3 were top-ranked in the High-PRS vs. Low-PRS comparison and also appeared among the top ten in the High-PRS vs. HC analysis. These findings implicate IRF7, BATF3, and other TFs as potential shared regulators of transcriptional activity in pDCs and monocytes in SLE patients with high polygenic risk.

Next, we performed protein–protein interaction network analysis of upregulated genes in pDCs and monocytes from high PRS patients (Figure 5D). In pDCs, ISG15 emerged as a central hub, interacting with STAT1, MX1, and USP18. Monocyte networks were more complex, with prominent STAT (STAT1/3/4), interleukin receptor (IL15RA, IL2RG, IL21R), and NF-κB (NFKBIA, NFKB2) clusters. Mapping IRF7s and BATF3s predicted targets onto the networks revealed substantial overlap, though distinct TF-specific modules were observed, e.g., STAT4 and IL15RA (BATF3), and HERC5 and TRIM25 (IRF7). These results suggest IRF7 and BATF3 as key regulators of inflammatory gene networks in SLE.

### 3.8. Comparison of PRS-Stratified SLE Signatures with Published Datasets

To assess how identified RNA expression profiles align with previously published transcriptomic studies in SLE, we performed public data enrichment analysis using the top 100 upregulated DEGs from the High-PRS and Low-PRS groups vs. HCs comparisons (Supplementary table S7). Among the top-ranked studies, 84% showed stronger enrichment for DEGs from the High-PRS group compared to those from the Low-PRS group. Interestingly, the analysis included several studies focusing on childhood onset SLE, supporting the importance of high genetic risk for early SLE onset.

## 4. Discussion

We observed a strong association between high PRS and an elevated IFN signature in patients with SLE, in remission or low clinical disease activity and ongoing HCQ treatment at adequate therapeutic concentration. In contrast, patients with low PRS demonstrated a weaker IFN signature, with IFN scores comparable to those of HCs. This divergence in IFN pathway activation is interesting and may at least partly explain the poorer prognosis reported in patients with high genetic risk[12]. It is also consistent with previous findings linking high baseline IFN signature to a higher risk of future SLE flares[23]. Importantly, our findings suggest that antimalarial therapy alone may be insufficient to fully suppress IFN-driven immune activation in patients with a high genetic burden, despite apparent clinical remission. These observations thus have potential implications for current treat-to-target strategies in SLE, as existing recommendations do not address the need of disease control at the molecular level in patients with high genetic risk. Thus, incorporating genetic risk stratification into clinical practice could help guide individualized long-term management. Further studies are needed to determine whether intensified maintenance therapy beyond HCQ can reduce flare risk and improve outcomes in this genetically high-risk subgroup.

Given the observed enrichment of both type I and type II IFN response pathways in our analysis, and the substantial overlap between genes induced by type I IFNs and IFN-γ[24], we generated a composite IFN score to capture the shared activation of these pathways. To disentangle their individual contributions, we also computed separate type I and type II IFN scores. Both were significantly elevated across multiple immune cell subsets in High-PRS patients compared to HCs, but not in Low-PRS vs. HCs comparisons. The increase was more pronounced for the type II IFN score. While this may seem unexpected, given the well-established role of type I IFNs in SLE[1] and limited therapeutic success targeting type II IFN[25], type II IFN signaling has been reported in SLE and may be more prominent in specific patient subsets or disease stages[26]. Additionally, HCQ, a standard SLE therapy, preferentially inhibits type I IFN signaling, potentially allowing type II IFN activity to persist despite treatment[27].

Our previous work has shown that individuals with higher genetic risk not only have an increased likelihood of developing SLE but also tend to exhibit a more severe clinical phenotype [12]. [12, 28]. Although the underlying mechanisms remain unclear, the cumulative effect of risk variants affecting immune regulation is a likely contributor. Supporting this, recent findings by Chung et al. showed that individuals with high PRS were exposed to lower ultraviolet light prior to disease onset compared to those with lower genetic risk[29], suggesting a lower threshold for disease activation in genetically susceptible individuals. Taken together, these observations support the notion that high genetic risk may lower the threshold for disease activation and relapse. In this study, we show that increased IFN signaling represents one mechanism by which genetic risk translates into heightened immune activation.

Given that pDCs are the primary producers of type I IFNs upon immune complex recognition and endosomal TLR7/9 ligation[30], we specifically investigated this cell subset. We observed significant enrichment of type I IFN signaling pathways in High-PRS pDCs patients compared to HCs, as well as between High-PRS and Low-PRS pDCs, indicating sustained IFN pathway activation in these cells among individuals with elevated genetic risk, despite treatment with HCQ. In contrast, genes unrelated to IFN signaling showed minimal pathway enrichment. This may reflect the low representation of pDCs in our dataset, consistent with their reduced presence in peripheral blood in SLE due to migration into inflamed tissues[31, 32]. Nonetheless, these observations align with the well-established role of pDCs, whose primary function upon activation is the robust production of type I IFNs[33, 34]. Together, our results suggest that genetic risk contributes to heightened IFN pathway activity in pDCs and that HCQ may be less effective at suppressing this signaling in patients with high PRS.

Similar to our findings in pDCs, we observed strong enrichment of IFN signaling pathways in monocytes from High-PRS patients compared to those from Low-PRS, suggesting that genetic risk modulates the intensity of IFN pathway activation in monocytes. However, monocytes from Low-PRS patients also demonstrated significant enrichment of IFN-related pathways relative to HCs, indicating that this cell type may exhibit a more broadly activated IFN signature in SLE, regardless of genetic risk. In addition to IFN signaling, several immune pathways were selectively enriched in the High-PRS group, including the TLR3 signaling cascade, which is known to mediate type I IFN induction in response to double-stranded RNA. Although TLR3 expression is typically low in monocytes at baseline, it can be upregulated by IFN-α/β, suggesting a potential feed-forward loop[35, 36]. These findings raise the possibility that, in SLE, and particularly in individuals with high genetic risk, type I IFN-driven upregulation of TLR3 may prime monocytes for heightened responsiveness to dsRNA, thereby amplifying their contribution to the type I IFN response. Thought speculative, this mechanism may represent an additional axis of dysregulated innate immunity in genetically susceptible patients.

Our analysis of TFs enriched in pDCs and monocytes identified IRF7 and BATF3 as shared regulators of differential gene expression in individuals with high polygenic risk for SLE. In pDCs, the robust and sustained production of type I IFNs is critically dependent on their constitutively high basal expression of IRF7 [37], a pattern confirmed by our single-cell transcriptomic data. This response is initiated through endosomal sensing of nucleic acid-containing immune-complexes, which activate a signaling cascade culminating in phosphorylation and dimerization of IRF7, enabling its nuclear translocation and transcriptional activation of type I IFN genes [38]. In contrast, monocytes express lower basal levels of TLR7, though its expression is rapidly upregulated following exposure to inflammatory stimuli, including type I IFNs [39]. Upon activation by pattern recognition receptors, IRF7 not only induces type I IFNs but also promotes the expression of pro-inflammatory cytokines that drive monocyte differentiation into inflammatory macrophages and dendritic cells, thereby amplifying tissue inflammation [40, 41]. Notably, monocytes carrying the SLE-risk *IRF7* haplotype exhibit a two-fold increase in IFN-α production following TLR stimulation[42, 43], aligning with our findings that implicate IRF7 as a central mediator of disease pathogenesis in genetically high-risk individuals.

BATF3, a basic leucine zipper transcription factor known for its role in classical DC differentiation, has also been implicated in IL-2 regulation and the development of tissue-resident CD4⁺ memory T cells in murine models of allergy as well as in inflammatory processes[44]. Additionally, BATF3 has been proposed to act as a transcriptional repressor of regulatory T cell differentiation[45], potentially modulating the balance between effector and regulatory immune responses. Although BATF3 is expressed in human monocytes and dendritic cells[46], its function in the context of SLE remains undefined, pointing to a previously uncharacterized regulatory pathway and a target for future investigation.

A key strength of our study is the integration of single-cell transcriptomic profiling with comprehensive genetic and clinical data in a large cohort of SLE patients and matched healthy controls. Notably, all patients were in remission and receiving only antimalarial therapy, minimizing confounding effects of active disease or immunosuppression. However, the study has limitations. Despite deep profiling of PBMCs, detection of rare cell populations or transcripts with low expression may have been constrained by technical sensitivity. Furthermore, the absence of longitudinal sampling precluded analysis of temporal dynamics, which could have provided additional insight into disease progression and treatment response.

### Conclusion

Our results demonstrate that individuals with a high polygenic risk for SLE exhibit persistent activation of interferon-related pathways despite achieving clinical remission and receiving antimalarial therapy. This observation supports the presence of a genetically driven immune activation state that is incompletely suppressed by conventional treatment. Together, these findings highlight the potential utility of genetic risk stratification in guiding therapeutic decision-making and emphasize the need for precision-based strategies to more effectively target subclinical immune dysregulation in SLE.

## Supporting information

Graphical abstract

## Data Availability

All data produced in the present study are available upon reasonable request to the authors.

## Acknowledgements

We thank Rezvan Kiani and Anna-Lena Åblad for collection of patient samples.

