## Supplementary tables 1-5, figures and methods for "Single-cell RNA-seq reveals a persistent interferon signature in immune cells from Systemic lupus erythematosus patients with high versus low polygenic risk scores despite antimalarial treatment"

**Supplementary file**

**Supplementary tables and figures**

| **Supplementary Table S1.** Clinical Characteristics of Patients with High and Low Polygenic Risk Scores (PRS) | | |
| --- | --- | --- |
| **Patient ID** | **PRS group** | **PRS score*** |
| P.PRS-H 1 | High | 9.00 |
| P.PRS-H 2 | High | 9.78 |
| P.PRS-H 3 | High | 9.35 |
| P.PRS-H 4 | High | 9.68 |
| P.PRS-H 5 | High | 10.21 |
| P.PRS-H 6 | High | 10.01 |
| P.PRS-H 7 | High | 10.72 |
| P.PRS-H 8 | High | 9.54 |
| P.PRS-H 9 | High | 10.93 |
| P.PRS-L 1 | Low | 7.56 |
| P.PRS-L 2 | Low | 8.04 |
| P.PRS-L 3 | Low | 6.75 |
| P.PRS-L 4 | Low | 6.76 |
| P.PRS-L 5 | Low | 7.03 |
| P.PRS-L 6 | Low | 6.90 |
| P.PRS-L 7 | Low | 8.08 |
| *PRS calculated according to Reid et al[1]. | | |

| Supplementary Table S2. The 57 SLE susceptibility SNPs included in the Polygenic Risk Score | | | | | |
| --- | --- | --- | --- | --- | --- |
| SNP | Gene | SLE OR | SNP | Gene | SLE OR |
| rs2476601 | *PTPN22* | 1.30 | rs849142 | *JAZF1* | 1.09 |
| rs1801274 | *FCGR2A* | 1.11 | rs4917014 | *IKZF1* | 1.28 |
| rs2205960 | *TNFSF4* | 1.34 | rs73366469 | *GTF2IRD1-GTF2I* | 1.73 |
| rs17849502 | *NCF2* | 2.16 | rs4728142 | *TNPO3-IRF5* | 1.55 |
| rs10911363 | *NCF2* | 1.13 | rs2070197 | *TNPO3-IRF5* | 1.86 |
| rs34889541^a^ | *CD45* | 1.15 | rs2980512^a^ | *FAM86B3P* | 1.11 |
| rs3024505 | *IL10* | 1.34 | rs2736340 | *BLK* | 1.28 |
| rs7579944 | *LBH* | 1.14 | rs7829816 | *LYN* | 1.05 |
| rs6740462 | *SPRED2* | 1.02 | rs1966115 | *PKIA-ZC2HC1A* | 1.07 |
| rs2111485 | *IFIH1* | 1.17 | rs877819 | *WDFY4* | 1.03 |
| rs10930046 | *IFIH1* | 1.07 | rs4963128 | *IRF7-PHRF1* | 1.31 |
| rs11889341 | *STAT4* | 1.68 | rs2732552 | *CD44* | 1.16 |
| rs6445972 | *ABHD6-PXK* | 1.06 | rs1308020^a^ | *RNASEH2C* | 1.15 |
| rs6445975 | *ABHD6-PXK* | 1.17 | rs3794060^a^ | *DHCR7-NADSYN1* | 1.02 |
| rs1132200 | *TMEM39A* | 1.27 | rs7941765^a^ | *ETS1-FLI1* | 1.13 |
| rs564799 | *IL12A* | 1.20 | rs10774625 | *SH2B3-ATXN2* | 1.18 |
| rs10936599 | *MYNN* | 1.04 | rs1059312 | *SLC15A4* | 1.30 |
| rs10028805 | *BANK1* | 1.21 | rs9652601 | *CLEC16A-CIITA-SOCS1* | 1.27 |
| rs907715 | *IL21* | 1.06 | rs34572943 | *ITGAM-ITGAX* | 1.51 |
| rs7726414 | *TCF7-SKP1* | 1.01 | rs223881 | *CCL22* | 1.16 |
| rs7708392^a^ | *TNIP1* | 1.32 | rs1170426^a^ | *ZPF90* | 1.17 |
| rs2431697 | *PTTG1-MIR146A* | 1.22 | rs2280381 | *IRF8* | 1.21 |
| rs17603856 | *ATXN1* | 1.06 | rs2941509 | *IKZF3* | 1.45 |
| rs11755393 | *UHRF1BP1* | 1.27 | rs930297 | *GRB2* | 1.00 |
| rs2762340 | *ANKS1A* | 1.07 | rs3093030 | *ICAM1-ICAM4-ICAM5* | 1.02 |
| rs597325 | *BACH2* | 1.08 | rs2304256 | *TYK2* | 1.36 |
| rs6568431 | *PRDM1-ATG5* | 1.17 | rs11697848 | *RNF114* | 1.02 |
| rs6932056 | *TNFAIP3* | 1.83 | rs7444 | *UBE2L3-YDJC-HIC2* | 1.22 |
| rs2327832 | *OLIG3-LOC100130476* | 1.20 |  |  |  |
| SNPs and corresponding odds ratios included in the non-HLA weighted polygenic risk score by Reid *et al* [1]. ^a^For SNPs not occurring on the ImmunoChip, the proxy SNP with the highest linkage disequilibrium[2] was used; for rs34889541, rs16843520 was used (r^2^=1.00); for rs7708392, rs6889239 was used (r^2^=0.99); for rs2980512, rs2948286 was used (r^2^=0.96); for rs1308020, rs489574 was used (r^2^=1.00); for rs3794060, rs4944062 was used (r^2^=1.00); for rs7941765, rs6590343 was used (r^2^=0.99); for rs1170426, rs1170427 was used (r^2^=1.00). | | | | | |

| **Supplementay Table S3**. Clinical characteristics of patients with high or low Polygenic Risk Score | | | |
| --- | --- | --- | --- |
|  | **High-PRS** | **Low-PRS** | **p-value** |
| **Demographic characteristics** |  |  |  |
| Female sex | 9 (100) | 7 (100) | - |
| Age at diagnosis | 26.4 ± 10.1 | 28.4 ± 12.3 | 0.74 |
| Age at sampling | 48.8 ±9.3 | 52.3 ±11.1 | 0.51 |
| Disease duration | 22.3 ± 6.1 | 23.9 ± 9.3 | 0.96 |
| ACR 1997 | 5.1 ± 1.4 | 6.0 ± 1.5 | 0.26 |
| ACR 1 Malar rash | 5 (56) | 3 (43) | 0.34 |
| ACR 2 Discoid rash | 2 (22) | 2 (29) | 0.42 |
| ACR 3 Photosensitivity | 5 (56) | 4 (57) | 0.39 |
| ACR 4 Oral ulcer | 2 (22) | 1 (14) | 0.45 |
| ACR 5 Arthritis | 6 (67) | 7 (100) | 0.15 |
| ACR 6 Serositis | 1 (11) | 3 (43) | 0.17 |
| ACR 7 Renal disorder | 2 (22) | 4 (57) | 0.16 |
| ACR 8 Neurological disorder | 1 (11) | 0 (0) | 0.56 |
| ACR 9 Hematological disorder | 6 (67) | 4 (57) | 0.37 |
| ACR 10 Immunologic disorder | 7 (78) | 7 (100) | 0.30 |
| ACR 11 ANA | 9 (100) | 7 (100) | - |
| SDI score | 0.22 ± 0.67 | 0.29 ± 0.76 | 0.93 |
| **At sampling** |  |  |  |
| SLAQ | 6.22 ± 4.11 | 7.71 ± 5.53 | 0.56 |
| SLEDAI-2K | 1.00 ± 1.00 | 1.14 ± 1.95 | 0.72 |
| LLDAS | 9 (100) | 7 (100) | - |
| Clinical SLEDAI-2K | 0.22 ± 0.67 | 0.29 ± 0.76 | 0.93 |
| Anti-dsDNA | 1 (11) | 2 (33) | 0.34 |
| Anti-SSA (Ro52) | 2 (22) | 2 (29) | 0.42 |
| Anti-SSA (Ro60) | 4 (44) | 2 (29) | 0.33 |
| Anti-SSB | 2 (22) | 1 (14) | 0.45 |
| Anti-RNP | 1 (11) | 0 | - |
| Anti-Sm | 0 | 0 | - |
| Low complement* | 2 (22) | 1(14) | 0.45 |
| HCQ treatment mg/week, ±SD | 1733 ±539 | 1833 ±557 | 0.71 |
| Serum HCQ, (ng/ml) ±SD** | 1461 ± 683.3 | 999.1 ± 496.8 | 0.22 |
| Serum IFN-α, mean (pg/ml) ±SD | 0.43±0.57 | 0.12±0.15 | 0.34 |
| Data are n (%) or mean ± SD. PRS, Polygenic Risk Score; SDI, Systemic Lupus Erythematosus Damage Index[3] ; ACR, American College of Rheumatology[4, 5]; ANA, antinuclear antibody; SLAQ, Systemic Lupus Erythematosus Activity Questionnaire[6]; SLEDAI-2K, Systemic Lupus Erythematosus Disease Activity Index[7]; LLDAS, Lupus Low Disease Activity State [8]; Clinical SLEDAI-2K, SLEDAI-2K excluding complement and dsDNA antibodies; *Low complement according to SLEDAI.. **Hydroxychloroquine (HCQ) measured in serum, one patient in the Low-PRS group was taking chloroquine phosphate and was not included in the analysis; IFN, interferon | | | |

| **Supplementary Table S4.** Immune cell subsets in patients with SLE stratified by SLE-polygenic risk score and healthy controls,  determined by flow cytometry of PBMCs | | | |
| --- | --- | --- | --- |
| Cell surface marker | High SLE- PRS | Low SLE- PRS | HCs |
|  | Mean± SD (%) | Mean± SD (%) | Mean± SD (%) |
| CD19 | 5.6±2.3 | 4.7±2.5 | 4.6±1.7 |
| CD3 | 35±9.4 | 47±11 | 51±8.8*** |
| CD4 | 58±14* | 56±19^**^ | 71±4.5*** |
| CD8 | 22±9.9* | 30.5±20^**^ | 24±3.2*** |
| CD14 | 23±4.9 | 18±4.3 | 23±7.8 |
| CD56 | 15±4.9 | 13±4.3 | 17±4.7 |
| BDCA2 | 0.38 ±0.17 | 0.53 ±0.21 | 0.67 ±0.23 |
| SLE: systemic lupus erythematosus, PRS: polygenic risk score,  PBMC: peripheral blood mononuclear cells, HC: healthy controls,  SD: standard deviation.  *Data available for 7 of 9 the individuals.  **Data available for 6 of7 the individuals.  *** Data available for 3-5 of 6 the individuals. | | | |

| **Supplementary Table S5.** Interferon stimulated gene (ISGs) sets used for IFN score calculation | | |
| --- | --- | --- |
| **Type I ISGs** | **Type II ISGs** | **Type I or II ISGs** |
| IFI27  IFI44L  USP18  CMPK2  RSAD2  IFIT3  IFI44  -  MX1  EPSTI1  OASL  -  -  ISG15  -  OAS1  EIF2AK2  -  IRF7  -  LY6E  ADAR  HERC6  PARP14  PARP9  DDX60 | IFI27  IFI44L  USP18  CMPK2  RSAD2  IFIT3  IFI44  IFIT1  MX1  EPSTI1  OASL  OAS3  NAMPT  ISG15  XAF1  -  EIF2AK2  NFKBIA  IRF7  TNFAIP3  LY6E | IFI27  IFI44L  USP18  CMPK2  RSAD2  IFIT3  IFI44  IFIT1  MX1  EPSTI1  OASL  OAS3  NAMPT  ISG15  XAF1  OAS1  EIF2AK2  NFKBIA  IRF7  TNFAIP3 |
| The table lists the top 20 differentially expressed genes (DEGs) used to calculate interferon (IFN) scores in single-cell RNA-seq analysis. Each column corresponds to a specific gene set: genes found in the Hallmark IFN-α list, genes found in the Hallmark IFN-γ list, and genes found in either the IFN-α or IFN-γ Hallmark lists. | | |

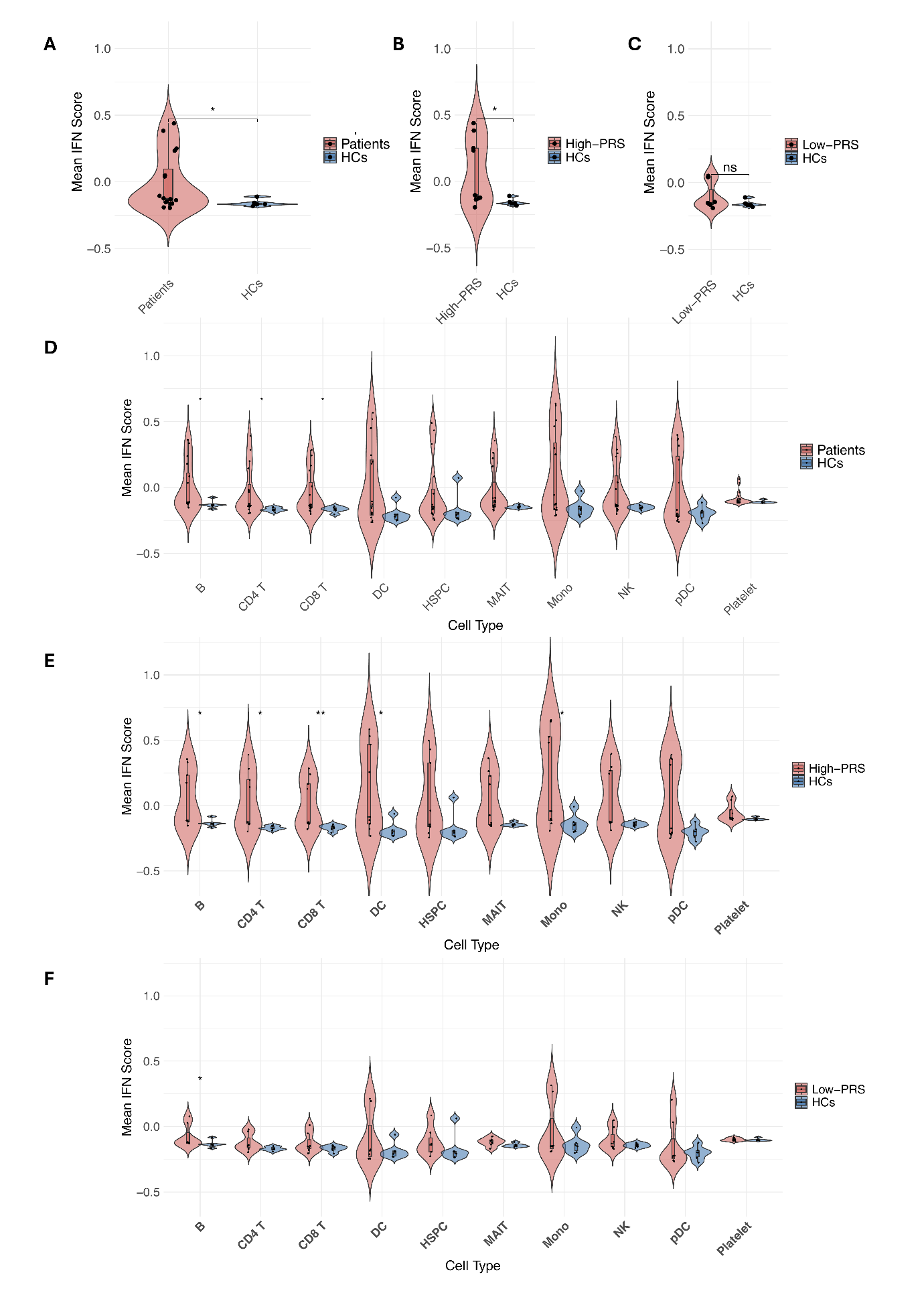

**Supplementary Figure S1. Type I interferon scores in patients and healthy controls**

Violin plots showing the distribution of type I interferon (IFN-α) response scores in patients and controls. Scores were computed per cell and averaged per sample. Panels A–C show pseudo-bulk comparisons (all cells combined) between all patients vs. healthy controls, High-PRS patients vs. healthy controls, and Low-PRS patients vs. healthy controls, respectively. Panels D–F show the same comparisons stratified by cell type. Statistical significance was assessed using the Mann–Whitney U test.

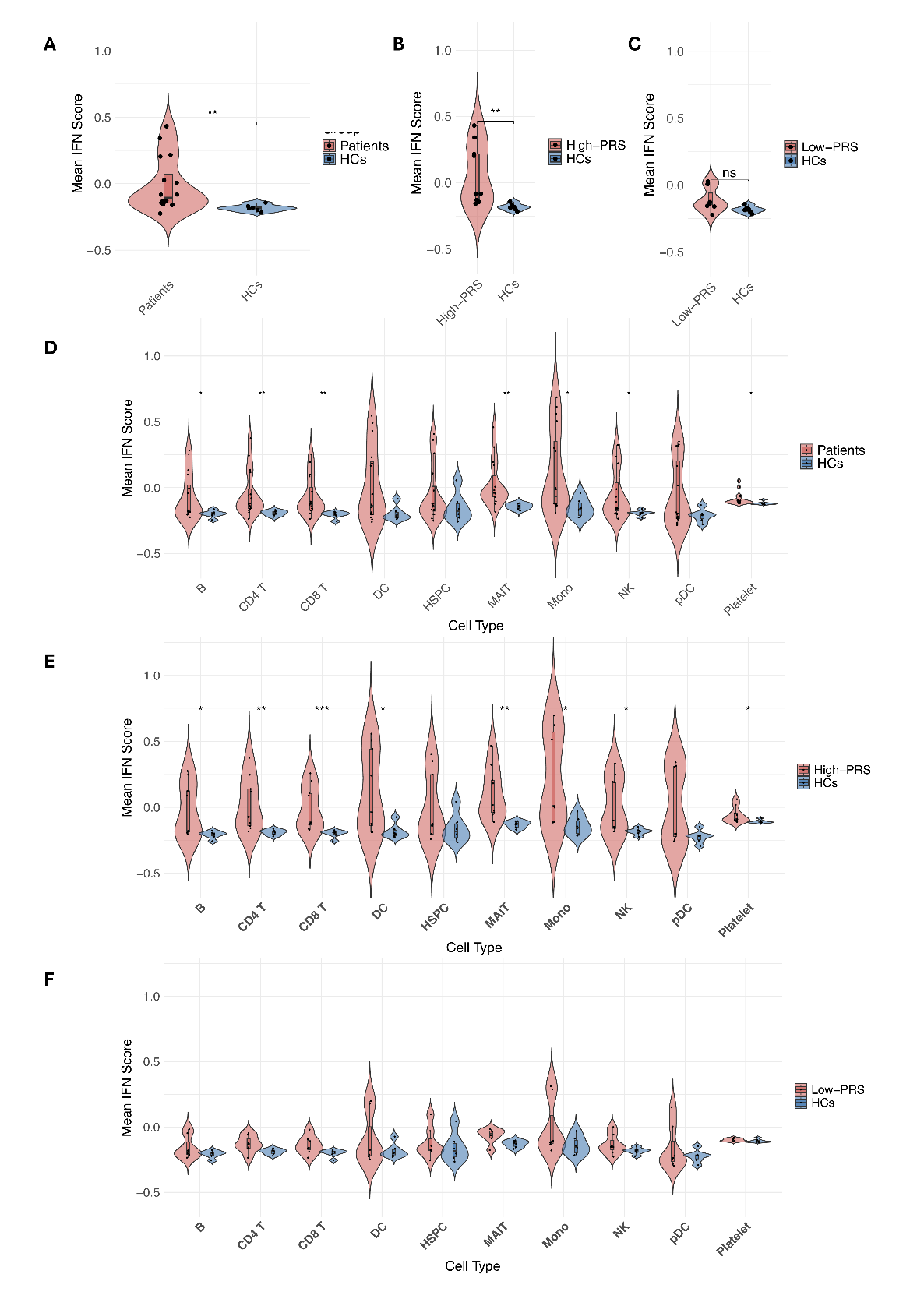

**Supplementary Figure S2. Type II interferon scores in patients and healthy controls**

Violin plots showing the distribution of type II interferon (IFN-γ) response scores in patients and controls. Scores were computed per cell and averaged per sample. Panels A–C show pseudo-bulk comparisons (all cells combined) between all patients vs. healthy controls, High-PRS patients vs. healthy controls, and Low-PRS patients vs. healthy controls, respectively. Panels D–F show the same comparisons stratified by cell type. Statistical significance was assessed using the Mann–Whitney U test.

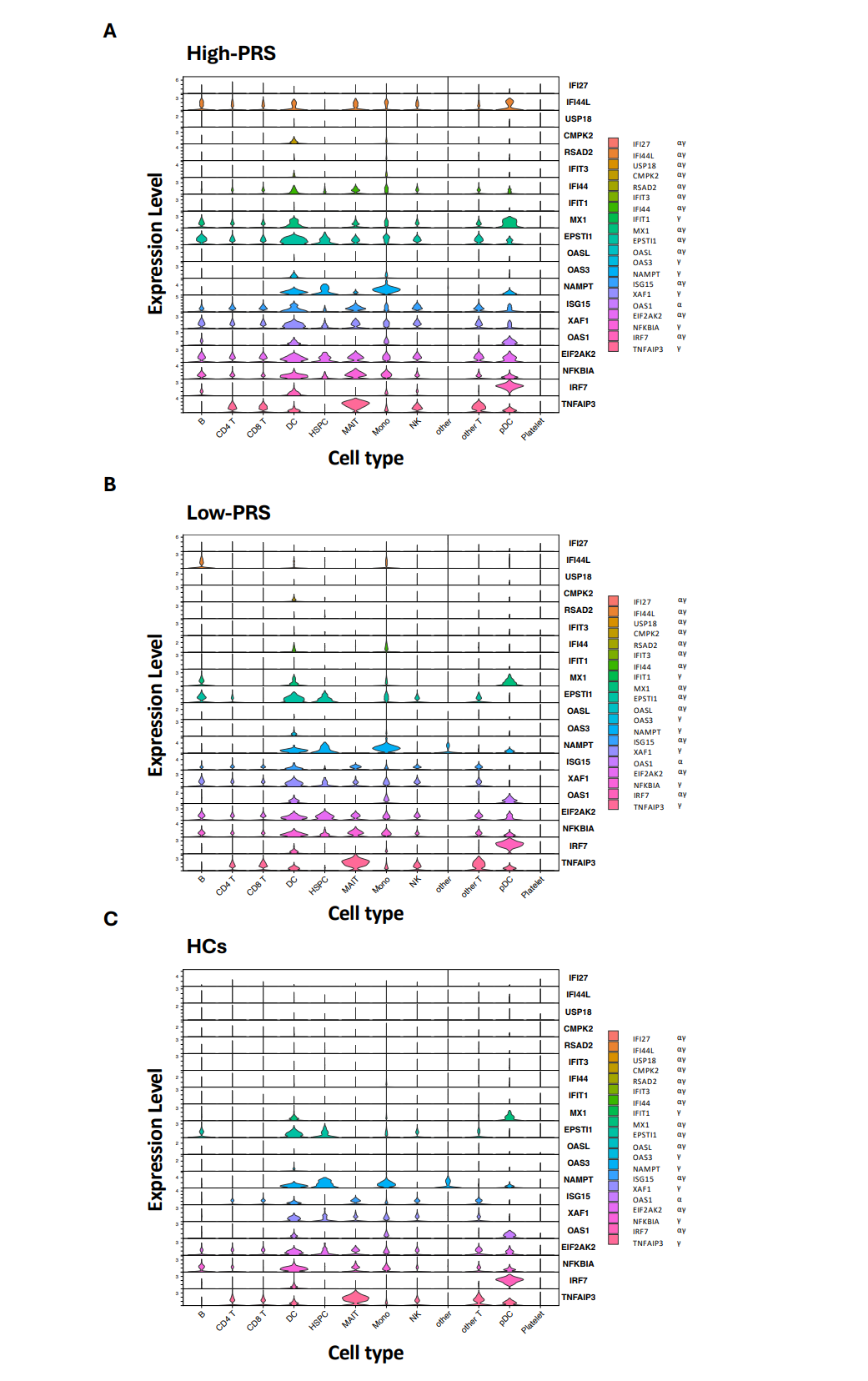

**Supplementary Figure S3. Expression of interferon-stimulated genes (ISGs) across cell types in High-PRS, Low-PRS, and Healthy Controls.** Stacked violin plots showing the expression of interferon (IFN)-stimulated genes (ISGs) previously used to calculate the IFN score, across immune cell types in (A) High-PRS, (B) Low-PRS and (C) Healthy Controls (HC).

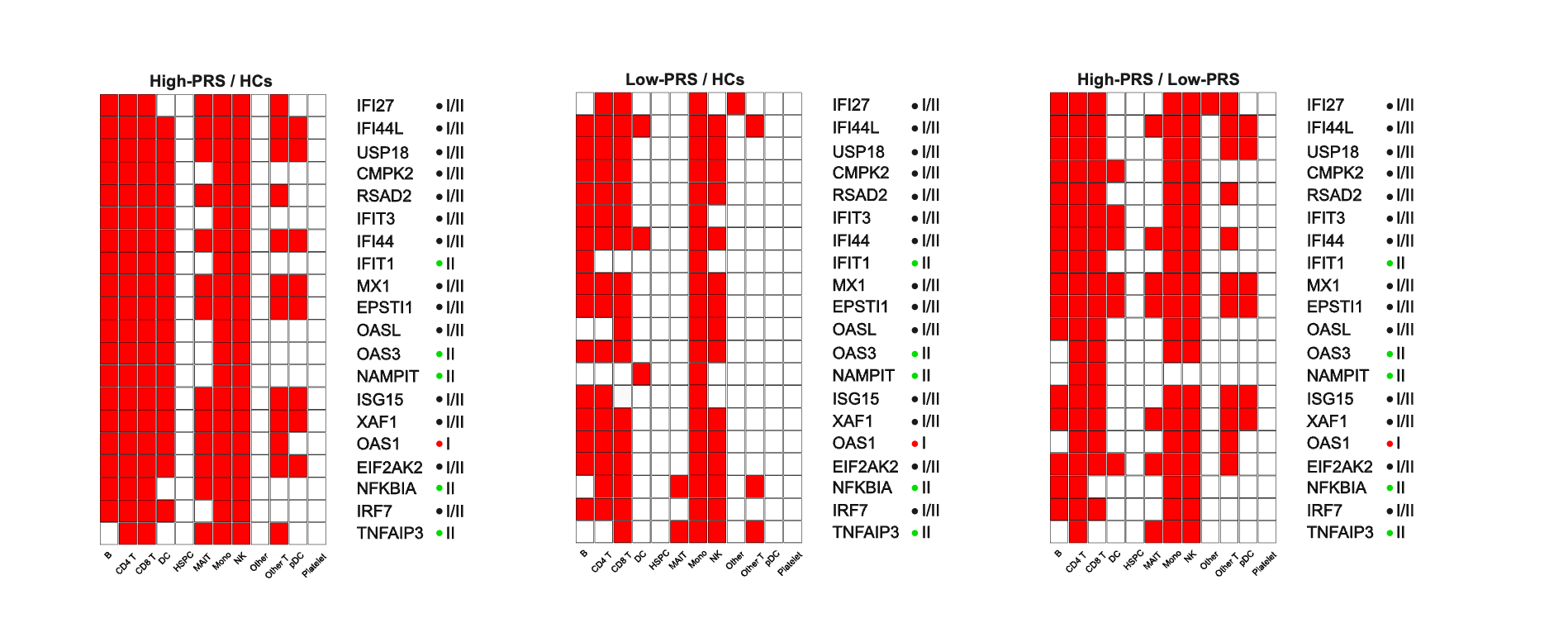

**Supplementary Figure S4. Differential expression of interferon (IFN)-stimulated genes** (ISGs), used for IFN score calculation the, across immune cell types (shown in the same order as in Figure S3). Differentially expressed genes (p_adj <0.05, magnitude of log2 fold-change > 0.25) in each cell type are shown as red rectangles.

**Supplementary Methods**

**Genotyping and quality control**

Genotyping was performed using the Illumina 200K Immunochip SNP array. Standard quality control (QC) procedures were applied to exclude low-quality samples and variants. Samples with a call rate below 95% and SNPs with call rate below 100% were excluded. To assess ancestry, principal component analysis (PCA) was conducted on the 1000 Genomes Project data. Samples positioned more than 5 standard deviations (SD) in each of the 5 principal components from the European cluster were excluded.

Sex discrepancies were evaluated using the inbreeding coefficient (F): samples annotated as male with F close to zero and females with F close to one were excluded. Samples showing abnormal autosomal heterozygosity rates, defined as more than 5 standard deviations from the mean Wright’s inbreeding coefficient (F), were also excluded. Cryptic relatedness was assessed using the kinship coefficients based on identity-by-descent (IBD). One individual from each pair of samples related up to the second degree was excluded.

SNPs with a minor allele frequency (MAF) < 1% or Hardy–Weinberg equilibrium (HWE) p-values within an FDR < 5% (based on controls only) were excluded, and to avoid falsely low cumulative genetic load estimates, only individuals with complete (100%) genotyping for all 57 SNPs were retained[9].

**Cell Type Annotation** Annotation of cell types was performed using the Azimuth reference-based mapping framework developed by the Satija lab, which projects single-cell query data onto a well-curated reference atlas to assign cell type labels[10, 11]. We utilized the PBMC reference dataset (https://azimuth.hubmapconsortium.org/references/human_pbmc/), which comprise approximately 263,000 PBMCs from healthy donors. The annotation was conducted via Seurat’s Azimuth workflow in R, and the predicted cell type identities were added into the Seurat object metadata for downstream differential expression and pathway analyses.

**Differential Gene Expression Analysis Across Cell Types**

Differential expression analyses were performed using Seurat. Comparisons were conducted both at the single-cell level and after pseudobulking (aggregating counts per sample) to capture patient-specific effects. Two types of comparisons were made: patients (High-PRS + Low-PRS) versus healthy controls (HCs), and separate comparisons of High-PRS versus HCs and Low-PRS versus HCs. Genes with an adjusted p-value < 0.05 and log2 fold change > 0.25 were considered upregulated, while genes with log2 fold change < -0.25 were considered downregulated.

**Heatmap of Top Differentially Expressed Genes**

To identify the top genes specifically upregulated in each group, differential expression analyses were performed by comparing each group against all others. Genes expressed in at least 25% of cells and exhibiting a magnitude of log2 fold-change > 0.25 were retained, considering only protein-coding genes. The resulting group-specific markers were used to generate heatmaps, with interferon-stimulated genes (ISGs) highlighted for visual emphasis.

For sample-level visualization, average expression per disease state was calculated using Seurat. The expression matrix was Z-score scaled across genes. Heatmaps were generated, applying hierarchical clustering to both genes and samples.

**Interferon Module Scoring and Visualization**

Interferon (IFN) response score was quantified based on the expression of interferon induced genes. These genes were identified by retrieving Hallmark Interferon Alpha Response and Hallmark Interferon Gamma Response gene sets from MSigDB[12]. Differentially expressed genes (adjusted p < 0.05, |log2FC| > 0.25) overlapping either interferon Hallmark set were ranked by p-value and fold change, and the top 20 genes were selected to define the IFN score. The interferon (IFN) score was calculated per cell using Seurat’s AddModuleScore function. Patient-level IFN scores were obtained by averaging IFN score values across all cells from each sample. Group comparisons were performed sequentially (Patients vs. HCs, High-PRS vs. HCs, and Low-PRS vs. HCs) using the Mann–Whitney U test. For each comparison, violin plots were generated to visualize bulk-level IFN score differences, followed by separate plots stratified by cell type. The same analysis was then repeated using two additional IFN scores derived from the new 20 gene sets: one restricted to genes intersecting with Hallmark Interferon Alpha Response and another restricted to genes intersecting with Hallmark Interferon Gamma Response. For both scores, all three comparisons (Patients vs. HCs, High-PRS vs. HCs, and Low-PRS vs. HCs) were evaluated at the bulk and cell-type levels using the same statistical and visualization approaches.

**Functional enrichment analysis**

For the enrichment analysis of known functional human pathways we used Reactome Gene Sets for the unranked lists of DEGs[9]. For monocytes and pDCs enrichment analysis was performed for all upregulated DEGs (monocytes: High-PRS vs. HCs (n=1,226), Low-PRS vs. HCs (n=901), High-PRS vs. Low-PRS (n=478); pDCs: High-PRS vs. HCs (n=53) and Low-PRS vs. HCs (n=9) and High-PRS vs. Low-PRS (n=22)). The analysis of non-IFN DEGs excluded DEGs known to be interferon-induced according to MSigDB signatures for Hallmark Interferon Response, alpha or gamma.

For limiting bias in enrichment analysis, a custom background group was used which included all genes, found to be expressed in PBMC samples in our study. Terms were estimated to be enriched if they passed Benjamini-Hochberg adjusted p-value tests, contained a minimum overlap of 2 genes for each pathway and enrichment factor (the ratio between the observed counts and the counts expected by chance) was higher than 1.2. For three group-based genes lists, enriched terms were prioritized based on their significance for each individual group and on their group relations (prioritizing the most common and the most group-specific pathways).

For identification of potential common upstream transcription factors (TFs), regulating DEGs, we performed TF enrichment analysis using the multiomic-based ChEA3 method[13]. For analysis we used the same lists of upregulated DEG lists for monocytes and pDCs as in the functional enrichment analysis. These genes were compared with ChEA3 libraries of TF target genes and the top 15 predicted common TFs were selected by the lowest MeanRank metric.

**Network analysis**

Protein–protein interaction (PPI) networks were constructed using STRING-DB, incorporating physical interaction data annotated from "experiments" and "databases" sources in STRING-DB. For the **plasmacytoid dendritic cell (pDC)** network, a medium-confidence interaction threshold (combined score ≥0.400) was applied. To enhance clarity in visualization, the **monocyte** network was restricted to high-confidence interactions only (combined score ≥0.900). Subnetworks displayed include only clusters of ≥4 directly connected proteins. Network plots highlight predicted transcriptional targets of **IRF7** and **BATF3**, based on target gene annotations from the ChEA3 transcription factor enrichment database.

**Preparation of peripheral mononuclear cells (PBMC) and flow cytometry**

Blood samples were collected in Vacutainer tubes (BD) containing EDTA as anticoagulant and PBMC were isolated by Ficoll-Paque (GE Healthcare) gradient centrifugation. Cell viability was ≥98% determined by Trypan blue staining.

An aliquot of the isolated PBMC were stained with fluorochrome labelled monoclonal antibodies to CD19, CD3, CD4, CD8, CD14, CD56 and BDCA-2 or with isotype controls (BD Bioscience or Miltenyi Biotec). Cells were acquired with a FACSCanto II instrument (BD Biosciences) and data was analysed with FlowJo (v.10.10.0.) software.
