## Supplementary figures and images for "Single-cell RNA-seq reveals a persistent interferon signature in immune cells from Systemic lupus erythematosus patients with high versus low polygenic risk scores despite antimalarial treatment"

### Graphical abstract

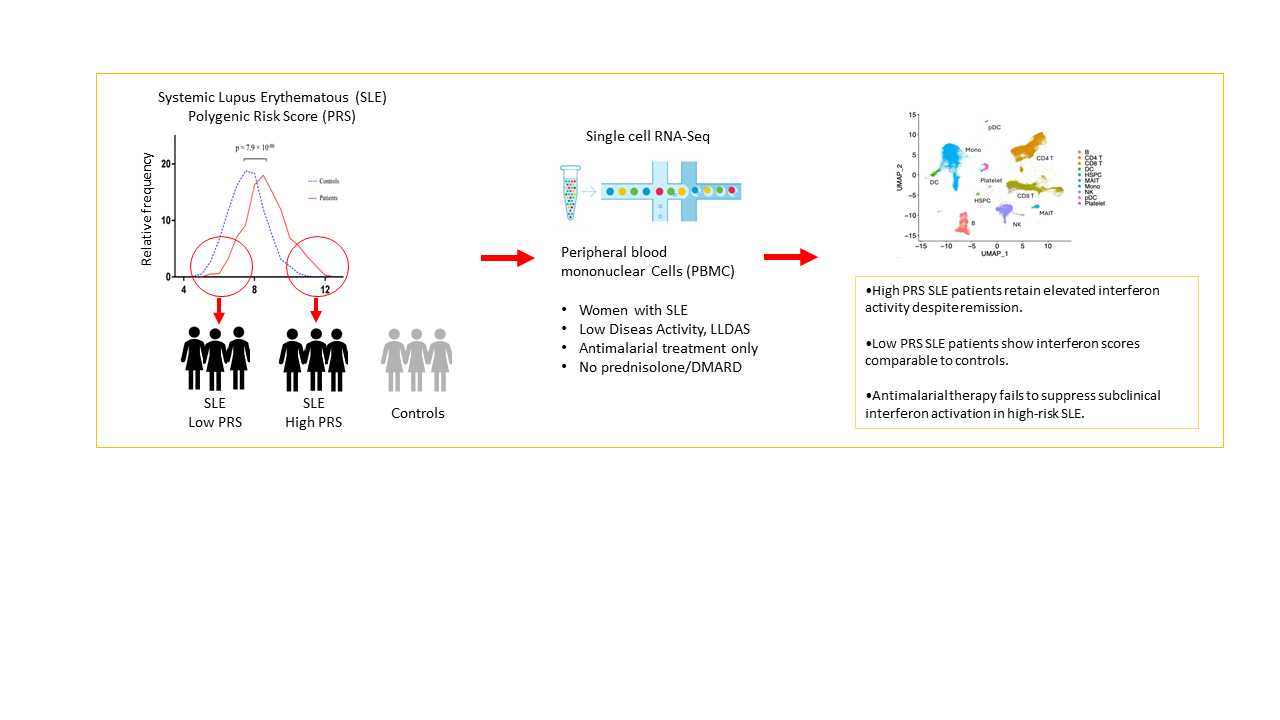
